# High-risk Human Papillomavirus (16, 18) are not the most common genotypes associated with cervical pre-cancer lesions: a retrospective study at a University Hospital in the Eastern-Province of Saudi Arabia

**DOI:** 10.1101/2020.03.17.20037465

**Authors:** Haitham Kussaibi, Reem Al Dossary, Ayesha Badar, Aroub Omar Muammar, Raghad Ibrahim Aljohani

**Author notes:** **Corresponding author: Dr. Haitham KUSSAIBI**, Assistant Professor, Department of Pathology, College of Medicine, Imam Abdulrahman Bin Faisal University (IAU), Dammam, Saudi Arabia.

## Abstract

**Objective:** High-risk HPV (human papillomavirus) is found to be responsible for 4.5% of all cancer, especially cervical cancer. The prevalence of high-risk HPV associated with cervical lesions is not well-known in Saudi Arabia. This study aims to highlight the genotypes of high-risk HPV associated with pre- malignant cervical lesions.

**Methods:** Over 6 years (2013 - 2018), 5091 Pap (Papanicolaou) smears results and 170 high-risk HPV test results were collected from the Information System at King Fahd University Hospital. Statistical analysis was performed using the software SPSS (Statistical Package for Social Sciences).

**Results:** Out of 5091 Pap smears, only 1.89% (n=96) were abnormal; 0.18% (n=9) were malignant (7 Squamous cell carcinomas and 2 adenocarcinomas), while 1.7% (n=87) showed pre-cancerous lesions, 44 ASCUS (Atypical Squamous Cells of Undetermined Significance), 17 LSIL (Low-grade Squamous Intraepithelial Lesions), 12 HSIL (High-grade Squamous Intraepithelial Lesions), and 14 AGC (Atypical Glandular Cells). Out of 170 patients co-tested for high-risk HPV, only 13.5% (n=23/170) had positive results (5 cases were positive for HPV16, 1 case was positive for both HPV16 and 18, while the remaining 17 cases were positive for high-risk HPV other than 16 or 18), among them, 6.47% (n=11/170) had normal Pap smear, while 7.06% (n=12/170) patients had abnormal Pap smear; 4 ASCUS, 6 LSIL and 2 HSIL. Statistical analysis showed a significant correlation between HPV findings and the Pap smear results (P-value 0.000), however, no significant correlation was found with the patients’ age and/or nationality.

**Discussion:** Our study showed a unique distribution of high-risk HPV genotypes which reflects different geographical infection patterns. Furthermore, the high association of high-risk HPV with normal Pap smears highlights the need, for all women at risk, to be co-investigated for high-risk HPV. These findings could help in customizing regional vaccine-combinations and screening programs.

## INTRODUCTION

Human papillomaviruses are a group of more than 100 viruses associated with a wide range of mucocutaneous manifestation ranging from benign warts, affecting multiple anatomical sites, to potentially malignant and malignant anogenital and oropharyngeal lesions [1]. HPV is found to be responsible for 4.5% of all cancer worldwide [2]. In addition to cervical cancer, which accounts for 83% of HPV associated cancer, HPV is also detected in penile, vulvar, vaginal, anal and oropharyngeal cancer [2].

The determinant of the progression of HPV associated lesions from benign to malignant lesions includes the HPV genotype, HPV viral load [3] and HPV persistence [4]. Up to date, 14 types of high-risk HPV (16, 18, 31, 33, 35, 39, 45, 51, 52, 56, 58, 59, 66 and 68) are known carcinogens [2], and persistence implies the detection of high-risk type for more than 6-12 months in cervical samples [5] since the majority of viral infections are cleaned in 1-1.5 year [3]. Studies on HPV viral load showed that the higher the viral load, the higher the progression to cancer will be [6].

Globally, the prevalence of HPV infection in women with normal Pap (Papanicolaou) smear is 11-12% [7, 8] and it varies with geographical location, and socioeconomic status with the highest percentage in the Caribbean (35.4 %) and lowest in Western Asia (1.7%). Furthermore, female age and degree of cervical pathological changes affect the prevalence of HPV detection. Of all HPV types, the most detected are HPV16 (3.2%), HPV18 (1.4%), HPV52 (0.9%), HPV31 (0.8%) and HPV58 (0.7%). Due-to the high burden of the virus and its associated high morbidity and mortality, prevention of HPV related cervical cancer, is implemented at multiple levels starting from vaccination, to screening to early diagnosis and management [9].

Currently, three vaccines are FDA (Food and Drug Administration) approved and they include 2, 4, and 9v (valent) vaccines; 2vHPV (Cervarix), 4vHPV (Gardasil), and 9vHPV (gardasil9) [10, 11]. HPV vaccines are made by recombinant DNA technology. The 2vHPV contains HPV16 and 18, and the addition of HPV6 and 11, the causative agent of an anogenital wart, makes the 4vHPVvaccine. While, the 9vHPV vaccine includes all types included in 4vHPV in addition to HPV 31, 33, 45, 52, and 58. These vaccines are recommended for females and males aged 11 - 26 years [11, 12].

For a screening purpose, a combination of cervical cytological evaluation (Pap smear) and DNA testing for high-risk HPV are currently employed and it is the preferred modalities to clinically segregate LSIL (Low-grade Squamous Intraepithelial Lesion) from the HSIL (High-grade Squamous Intraepithelial Lesion) and SCC (squamous cell carcinoma) [13]. Clinical outcome of patients having ASCUS (Atypical Squamous Cells of Undetermined Significance) and LSIL is documented to be similar and their treatment options are controversial [14]. Seventy percent of ASCUS and LISL show spontaneous regression [15]. ASCUS or LSIL progressing to HSIL is still debatable and yet remains a diagnostic and clinical challenge for cytopathologists, clinicians and patients. Performing colposcopy and biopsy for all such patients will markedly increase the rates of these procedures with a negative impact of patient’s physical and psychological health, especially for those patients whose lesions are not prone to develop into HSIL. Consequently, an additional triage comprising high-risk HPV PCR (Polymerase chain reaction) detection is recommended for all women with ASCUS/LSIL [16, 17].

High-risk HPV DNA testing has been shown in trials to be more sensitive than cervical cytology in the detection of intraepithelial lesions [18]. This increased sensitivity can safely lead to more spaced apart screening time intervals [19-21].

The prevalence and genotype of high-risk HPV infection detected in women with normal or abnormal cytology; ASCUS, LSIL, and HSIL) are so far unknown (according to “HPV and related diseases report” in Saudi Arabia published by the “HPV information center” on December 2018) [22, 23]. Our study aims to highlight the prevalence and genotype distribution of high-risk HPV infection and its association with cervical cancer and precancerous lesions in the Eastern-Province and other areas in Saudi Arabia.

## METHODS

### Study design

Retrospective cross-sectional study to determine the prevalence and genotype distribution of high-risk HPV and its association with cervical cancer and precancerous lesions in patients attending gynecology clinic at King Fahd University Hospital in the Eastern Province, Saudi Arabia for 6 years period (2013-2018). For routine cervical cancer screening, Pap smear is done for all married women aged 21-65 years, every 3 years without or with high-risk HPV molecular testing (for patients at elevated risk). Furthermore, high-risk HPV molecular testing is done for all patients showing ASCUS on cytological examination as per CAP (College of American Pathologist) recommendation.

**Ethical approval** was obtained from the SCRELC (Standing Committee on Research Ethics on Living Creatures) at Imam Abdulrahman Bin Faisal University with IRB-2018-01-157

### Cytological examination

Cervical Pap smears were collected at obstetrics and gynecology clinics in a commercially available liquid fixative vial (“SurePath” liquid-based Pap) to be processed in a semi-automated machine (The “PrepStain” System) which is a liquid-based thin-layer cell preparation processor. The “PrepStain” system produces the “SurePath” slides for microscopic examination by a pathologist and reported according to “The Bethesda System”.

### High-risk HPV detection

As our laboratory follows CAP (College of American Pathologist) standards, all cervical samples reported ASCUS on Pap smear, should be sent for high-risk HPV test. Furthermore, patients at risk are also investigated for high-risk HPV, in parallel to Pap smear. In both ways, Samples were preserved using the same vial “SurePath liquid-based Pap” mentioned above. Detection of high-risk HPV was done using “Xpert HPV” assay which is a fully automated multiplex qualitative real-time PCR using “Genexpert” by “CEPHEID”. This test detects and identifies all the 14 high-risk HPV types. The results are reported as negative or positive (HPV16, HPV18 and/or other high-risk HPV types without further specification).

## Statistical analysis

All Statistical analysis was performed using the software SPSS (Statistical Package for the Social Sciences) with a significant correlation on P-value = 0.05 or less.

## RESULTS

### 1. Prevalence of pre-cancerous and cancerous cervical lesions in women attending gynecology clinic at King Fahd University Hospital in the Eastern Province of Saudi Arabia (between 2013-2018)

Over the 6 years study period, 5091 Pap smears were tested, only 1.89% (n=96) were abnormal, and the remaining 4995 were normal (Table 1). Out of the 96 abnormal Pap smears, 0.18% (n=9) were malignant (7 Squamous Cell Carcinomas and 2 Adenocarcinomas) and 1.7% (n=87) showed pre-cancerous cervical lesions in the form of ASCUS, LSIL, HSIL, or AGC (Table 1).

**Table 1:** Pap smear results for women attending gynecology clinic, King Fahd University Hospital between (2013-2018):

| Pap smear results | Number | Percentage (%) |
| --- | --- | --- |
| ASCUS | 44 | 0.86 |
| LSIL | 17 | 0.33 |
| HSIL | 12 | 0.24 |
| AGC | 14 | 0.27 |
| Squamous cell carcinoma | 7 | 0.14 |
| Adenocarcinoma | 2 | 0.04 |
| Normal | 4995 | 98.11 |
| **Total Pap smear** | **5091** | **100** |

Among all 96 patients with abnormal Pap smears, only 31.25% (n=30) patients underwent cervical histology examination, among them, only 15 patients showed abnormal histology (malignant or pre- malignant), the remaining patients had either cervicitis or benign polyps by histological examination.

### 2. Results of patients co-tested for high-risk HPV genotypes (between 2013-2018)

Over the 6 years study period, only 170 women were co-investigated for high-risk HPV genotypes in addition to Pap smear (patients at high risk of infection and those who had ASCUS on Pap smear). Out of them, only 13.5% (n=23) cases were positive for high-risk HPV, while the remaining 86.5% (n=147) cases were negative (high-risk HPV not detected). Among the 23 patients positive for high-risk HPV, the most detected genotypes were “high-risk HPV other than 16 or 18” in 73.91% (n=17) patients, followed by HPV16 in 26.09% (n=6) patients, and finally HPV18 in 4.35% (n=1) patient. Almost, a similar distribution of high-risk HPV types was seen in patients with normal and abnormal Pap smear. Among the 86.5% (n=147) patients with negative high-risk HPV, the majority 70.59 % (n=120) had normal Pap smear, while the remaining 15.88% (n=27) had abnormal Pap smear (6 LSIL and 21 ASCUS). On the other hand, among the 13.5% (n=23) patients positive for high-risk HPV; 6.47% (n=11) had normal Pap smear, while 7.06% (n=12) patients had abnormal Pap smear; 4 ASCUS, 6 LSIL and 2 HSIL (Table 2).

### 3. Correlation of age, nationality and cytology examination with HPV status

There was a significant correlation between Pap smears findings and high-risk HPV status and vice versa (P-value 0.000).

**Table 2:** Results of the 170 patients co-tested for Pap smear and high-risk HPV:

| High-risk HPV genotyped (170 patients) | Normal Pap smear 77.07% (n=131/170) | Abnormal Pap smear 22.93% (n=39/170) |  |  |
| --- | --- | --- | --- | --- |
|  |  | ASCUS (n=25) | LSIL (n=12) | HSIL (n=2) |
| Not detected 86.5% (n=147) | 81.6% (n=120/147) | 18.3% (n=27/147) |  |  |
|  |  | 21 | 6 | 0 |
| Total detected 13.5% (n=23) | 47.8% (n=11/23) | 52.2% (n=12/23) |  |  |
|  |  | 4 | 6 | 2 |
| HPV16 (5 cases) | 3 | 0 | 2 | 0 |
| HPV 16 and 18 (1 case) | 0 | 0 | 1 | 0 |
| Other High-risk HPV (17 cases) | 8 | 4 | 3 | 2 |

Concerning the age distribution, our data showed that all the HPV16 and/or HPV18 patients were younger than 40 years old, while, the cases positive for high-risk HPV other than 16 or18 showed wide age distribution (21 - >61), however, there was no significant correlation between high-risk HPV status and age.

Considering that most of our patients, in the study, are Saudis, there was no significant correlation between high-risk HPV status and patients’ nationalities.

## DISCUSSION

This 6-year retrospective study conducted to disclose the association of different high-risk HPV types with abnormal Pap smears (pre-cancerous and cancerous cervical lesions), included a total of 5091 cervical Pap smears, the primary cervical cancer screening tool used in our center. Only 1.89% (n=96) Pap smears were abnormal. On the other hand, only 170 patients were co-tested for high-risk HPV in addition to Pap smear, among them, only 22.94% (n=39/170) had abnormal Pap smear 14.70% (n=25/170) diagnosed as ASCUS, 7.05% (n=12/170) as LSIL and 1.17% (n=2/170) as HSIL. Among those 22.94% (n=39/170) patients with abnormal Pap smear, high-risk HPV was detected in merely 7.05% (n=12/170) patients including 2.35% (n=4/170) had ASCUS, 3.53% (n=6/170) had LSIL and 1.18% (n=2/170) had HSIL. The remaining 15.88% (n=27/170) patients with abnormal Pap smear showed negative results for high-risk HPV (Table 2). HPV16, which is the most common type seen in ASCUS in many studies, has not been detected in any of the 25 cases of ASCUS in our study, however, only 16.66% (n=4/25) of ASCUS cases were positive for high-risk HPV other than 16 or 18. In an Iranian study conducted by Karimi-Zarchi et al in 2015, out of 180 cases of ASCUS evaluated, an overall high-risk HPV was seen in 58 (32.22%) with HPV16 seen in 46 (25.5%). This was followed by HPV18 in 2 (1.11%) and other high-risk HPV in 10 (5.5%) [24]. A study by Clifford et al, documented HPV16 in ASCUS to be 31% [25]. A Turkish study reported it to be 35% (n=33/94)[26]. Besides the variation in the types of HPV, the frequency of HPV positivity in ASCUS was also in a much lower range in our study than in other reported studies. These variations are highly reflective of different geographical representations of infection patterns. These also warrant a more elaborate and in-depth analysis of infection demographics, so that specific screening and follow up strategies could be developed, tailored to the endogenous regional needs which are so far highly deficient in this region.

In our study, a total of 12 cases of LSIL retrieved, 50% (n=6/12) were positive for high-risk HPV. This is in stark contrast to a Chinese study by Zheng et al. who reported 75.8% of women with LSIL to be high-risk HPV positive with HPV positivity rates declining with increasing age except in patients older than 60 years [27]. A Turkish study reported it to be 62% (n=18/29) [26]. Regarding HSIL, from an overall 12 cases, 16.66% (n=2) were seen to be high-risk HPV positive. In the Turkish study, it was reported to be 83% (n=5/6) [26].

Again, the local figures for the frequency of high-risk HPV in LSIL and HSIL show marked fluctuations from other international studies. Our limited sample size raises two issues. First, in our center, we do not have an organized, implemented screening program so most of the cases coming to the Gynecological clinic, were not tested for high-risk HPV, and these figures could be just reflecting an iceberg with completely unfathomed depths and boundaries. This highlights the need for a more vigilant screening program so the missed cases can be picked up and a true reflection of infection characteristics gets unraveled. Additionally, as pointed out earlier too, the frequency pattern with its limitation is still highlighting our regional infection characteristics.

High-risk HPV was not detected in 84% (n=21/25) of ASCUS cases. This could be attributed to the overdiagnosis of ASCUS or due to the negative predictive value of high-risk HPV testing. The acceptable range by CAP (College of American Pathologists) is only 50% of ASCUS cases can be negative for HPV.

Regarding ASC (Atypical Squamous Cells) overdiagnosis, this diagnostic entity is yet controversial to the end-users, the pathologists and clinicians, with no accompanying definite histological cervical lesion. It just signifies that the cervix might or might not have an associated pathology. Bethesda 2016 Committee recommended strict criteria for the ASC (Atypical Squamous cells) category, however, its overuse still high in many laboratories. Gupta et al reported ASC (Atypical Squamous cells) cytological diagnosis followed by a negative cervical biopsy diagnosis in 199 cases. They reviewed the initial cytological diagnosis and concluded that an initial over-diagnosis of ASC (Atypical Squamous cells) could be attributed to the presence of perimenopausal cells (17.6%), atypia attributable to reproductive tract infection in (14.6%) smears, hormonal induced alterations (8.5%), drying artifacts (3.5%) and immature metaplastic cell (2%) [28]. Many of these cases are not associated with significant lesions, which leads to many unnecessary colposcopies, biopsies, HPV typing or even repeated Pap tests.

Many studies showed that the majority of LSIL is associated with evidence of HPV infection (55% to 89% of LSIL are positive for high-risk HPV). However, in our study, negative high-risk HPV was seen in 50% (n=6/12) of LSIL cases. Barron et al reported a follow up of 468 patients from a total of 608 patients who had an initial diagnosis of LSIL with negative HPV [29]. HSIL was seen in 3% (n=14) and LSIL in 39.3% (n=184). No case of cervical carcinoma was seen. Bosquet et al in a retrospective study conducted in a period from 2003 to 2015 constituting 609 cases, reported that 12% of HSIL was negative for HPV [30]. In our setup, a stricter follow up of LSIL cases with negative high-risk HPV needs to be developed.

Out of a total of 170 cervical cytological cases tested for high-risk HPV, a major proportion comprising 86.47% (n=147/170) cases were negative for high-risk HPV. These negative cases combined with normal cytology were seen in 70.59% (n=120/170) cases.

Regarding high-risk HPV positive cases with normal cytology, it was detected in 6.4% (n=11/170). This prevalence is much higher than what has been reported in a regional Kuwaiti study in which 2.3 % (n=71/3011) of cases showed high-risk HPV positivity in normal cervical smears [31]. A study in Asia and Europe showed the variability of normal Pap smear associated with HPV infection, ranging from 3.8% in Thailand, 2.9% in Spain, to 1.6% in Vietnam [29]. A large cohort Dutch study (Population-Based Screening Study Amsterdam) reported it to be 3.8 %. However, our prevalence is far less than documented in a Turkish study which stated it to be 17.41% (n=35/201) [26]. Meta-analysis (PCR based) worldwide studies showed that HPV in patients with normal Pap smear is 10% [32]. Our prevalence pattern of 6.4% high-risk HPV in normal cervical smears is reflective of indigenous regional infection prevalence characteristics which have so far not been explored and fathomed. Our sample size in which the high-risk HPV testing was done was small but still, it forms a nidus on which more extensive regional studies can be based.

The presence of high-risk HPV in normal cervical smears highlights the need for an active screening program. A patient ≥30 years with a positive HPV and negative cytology is at higher risk of cervical cancer. The management of this category may include repeat HPV and cytology co-testing at 12 months. If the patient is still positive for HPV, colposcopy needs to be performed immediately [26].

A decreased prevalence of high-risk HPV in cases of ASCUS, LSIL, and HSIL in our set of cases along with a complete absence of HPV16 and 18 in ASCUS is in stark contrast to other regional and international studies. This variation is highly reflective of different geographical representations of infection patterns. Additionally, the presence of high-risk HPV in cases with normal cytological smears highlights the need for an active screening program tailored specifically to our endogenous, local, regional needs.

## Data Availability

Data are available upon a reasonable request

## Acknowledgments

This work was funded By DSR (deanship of scientific research) at Imam Abdulrahman Bin Faisal University (Project number 201815).

The authors are grateful to Dr. Ahlam Alghamdi, Consultant Gynecology and Mrs. Raeda Alkhateeb, Cytopathology Specialist at King Fahd University Hospital, for their valuable contribution.

## Conflict of Interest

The authors declare no conflict of interest in the paper.

Highlights, outlining the key findings and impact of the study

- HPV16 and 18, both represent 26% of all high-risk HPV detected (for vaccine customization).
- 50% of patients with positive high-risk HPV, showed normal Pap smear (add HPV test in screening).
- Only 16% of ASCUS cases and 50% of LSIL, were positive for high-risk HPV (diagnostic accuracy).

## REFERENCES

1. Serrano B, Brotons M, Bosch FX, Bruni L. Epidemiology and burden of HPV-related disease. Best practice & research Clinical obstetrics & gynaecology. 2018;47:14–26.

2. de Martel C, Plummer M, Vignat J, Franceschi S. Worldwide burden of cancer attributable to HPV by site, country and HPV type. I nt J Cancer. 2017;141(4):664–70.

3. de Sanjose S, Brotons M, Pavon MA. The natural history of human papillomavirus infection. Best practice & research Clinical obstetrics & gynaecology. 2018;47:2–13.

4. Crow JM. HPV: The global burden. Nature. 2012;488(7413):S2–3.

5. Syrjanen K. Persistent high-risk human papillomavirus (HPV) infections as surrogate endpoints of progressive cervical disease. Potential new endpoint for efficacy studies with new-generation (non-HPV 16/18) prophylactic HPV vaccines. European journal of gynaecological oncology. 2011;32(1):17–33.

6. Zhao X, Zhao S, Hu S, Zhao K, Zhang Q, Zhang X, et al. Role of Human Papillomavirus DNA Load in Predicting the Long-term Risk of Cervical Cancer: A 15-Year Prospective Cohort Study in China. T he Journal of infectious diseases. 2019;219(2):215–22.

7. Forman D, de Martel C, Lacey CJ, Soerjomataram I, Lortet-Tieulent J, Bruni L, et al. Global burden of human papillomavirus and related diseases. Vaccine. 2012;30 Suppl 5:F12–23.

8. Keinan Boker L, Twig G, Klaitman-Meir V, Derazne E, Shina A, Levine H, et al. Adolescent characteristics and incidence of pre-malignant disease and invasive tumors of the cervix. Int J Gynecol Cancer. 2020.

9. Gultekin M, Ramirez PT, Broutet N, Hutubessy R. World Health Organization call for action to eliminate cervical cancer globally. Int J Gynecol Cancer. 2020.

10. de Oliveira CM, Fregnani J, Villa LL. HPV Vaccine: Updates and Highlights. A cta cytologica. 2019;63(2):159–68.

11. Markowitz LE, Dunne EF, Saraiya M, Chesson HW, Curtis CR, Gee J, et al. Human papillomavirus vaccination: recommendations of the Advisory Committee on Immunization Practices (ACIP). MMWR Recommendations and reports : Morbidity and mortality weekly report Recommendations and reports. 2014;63(Rr-05):1–30.

12. Petrosky E, Bocchini JA, Jr., Hariri S, Chesson H, Curtis CR, Saraiya M, et al. Use of 9-valent human papillomavirus (HPV) vaccine: updated HPV vaccination recommendations of the advisory committee on immunization practices. M MWR Morb Mortal Wkly Rep. 2015;64(11):300–4.

13. Li T, Li Y, Yang GX, Shi P, Sun XY, Yang Y, et al. Diagnostic value of combination of HPV testing and cytology as compared to isolated cytology in screening cervical cancer: A meta-analysis. Journal of cancer research and therapeutics. 2016;12(1):283–9.

14. Katki HA, Schiffman M, Castle PE, Fetterman B, Poitras NE, Lorey T, et al. Five-year risks of CIN 2+ and CIN 3+ among women with HPV-positive and HPV-negative LSIL Pap results. Journal of lower genital tract disease. 2013;17(5 Suppl 1):S43–9.

15. Alanen KW, Elit LM, Molinaro PA, McLachlin CM. Assessment of cytologic follow-up as the recommended management for patients with atypical squamous cells of undetermined significance or low grade squamous intraepithelial lesions. Cancer. 1998;84(1):5–10.

16. Arbyn M, Ronco G, Anttila A, Meijer CJ, Poljak M, Ogilvie G, et al. Evidence regarding human papillomavirus testing in secondary prevention of cervical cancer. Vaccine. 2012;30 Suppl 5:F88–99.

17. Jordan J, Arbyn M, Martin-Hirsch P, Schenck U, Baldauf JJ, Da Silva D, et al. European guidelines for quality assurance in cervical cancer screening: recommendations for clinical management of abnormal cervical cytology, part 1. Cytopathology : official journal of the British Society for Clinical Cytology. 2008;19(6):342–54.

18. Ronco G, Dillner J, Elfstrom KM, Tunesi S, Snijders PJ, Arbyn M, et al. Efficacy of HPV-based screening for prevention of invasive cervical cancer: follow-up of four European randomised controlled trials. Lancet (London, England). 2014;383(9916):524–32.

19. Kitchener HC, Gilham C, Sargent A, Bailey A, Albrow R, Roberts C, et al. A comparison of HPV DNA testing and liquid based cytology over three rounds of primary cervical screening: extended follow up in the ARTISTIC trial. European journal of cancer (Oxford, England : 1990). 2011;47(6):864–71.

20. Madom LM, Boardman LA. HPV testing and cervical cancer screening: recommendations and practice patterns. Medicine and health, Rhode Island. 2005;88(10):362–3.

21. Patnick J. Review of recommendations on cervical cancer screening in the European Union. Minerva ginecologica. 2003;55(4):293–5.

22. Anfinan NM. Physician’s knowledge and opinions on human papillomavirus vaccination: a cross-sectional study, Saudi Arabia. B MC Health Serv Res. 2019;19(1):963.

23. Mousa M, Al-Amri SS, Degnah AA, Tolah AM, Abduljabbar HH, Oraif AM, et al. Prevalence of human papillomavirus in Jeddah, Saudi Arabia. A nn Saudi Med. 2019;39(6):403–9.

24. Karimi-Zarchi M, Tabatabaie A, Dehghani-Firoozabadi A, Shamsi F, Baghianimoghaddam M, Dargahi M, et al. The Most Common Type of HPV in Women with Atypical Squamous Cell of Undetermined Significance (ASCUS) in Pap Smear in Iran-Yazd. I nt J Biomed Sci. 2015;11(4):173–5.

25. Clifford GM, Smith JS, Plummer M, Muñoz N, Franceschi S. Human papillomavirus types in invasive cervical cancer worldwide: a meta-analysis. B r J Cancer. 2003;88(1):63–73.

26. Beyazit F, Silan F, Gencer M, Aydin B, Paksoy B, Unsal MA, et al. The prevelance of human papillomavirus (HPV) genotypes detected by PCR in women with normal and abnormal cervico-vaginal cytology. Ginekol Pol. 2018;89(2):62–7.

27. Zheng B, Yang H, Li Z, Wei G, You J, Liang X, et al. HPV test results and histological follow-up results of patients with LSIL Cervical Cytology from the Largest CAP-certified laboratory in China. J Cancer. 2017;8(13):2436–41.

28. Gupta S, Sodhani P. Reducing “atypical squamous cells” overdiagnosis on cervicovaginal smears by diligent cytology screening. Diagn Cytopathol. 2012;40(9):764–9.

29. Barron S, Austin RM, Li Z, Zhao C. Follow-up outcomes in a large cohort of patients with HPV-negative LSIL cervical screening test results. A m J Clin Pathol. 2015;143(4):485–91.

30. González-Bosquet E, Fernandez S, Sabra S, Lailla JM. Negative HPV testing among patients with biopsy-proven cervical intraepithelial neoplasia grade 2/3 or cervical cancer. I nt J Gynaecol Obstet. 2017;136(2):229–31.

31. Al-Awadhi R, Chehadeh W, Kapila K. Prevalence of human papillomavirus among women with normal cervical cytology in Kuwait. J Med Virol. 2011;83(3):453–60.

32. de Sanjosé S, Diaz M, Castellsagué X, Clifford G, Bruni L, Muñoz N, et al. Worldwide prevalence and genotype distribution of cervical human papillomavirus DNA in women with normal cytology: a meta-analysis. Lancet Infect Dis. 2007;7(7):453–9.

